# Altered thalamocortical structural connectivity in persons with schizophrenia and healthy siblings

**DOI:** 10.1101/2020.01.26.20018796

**Authors:** Beier Yao, Sebastiaan F. W. Neggers, René S. Kahn, Katharine N. Thakkar

## Abstract

Schizophrenia has long been framed as a disorder of altered brain connectivity, with dysfunction in thalamocortical circuity potentially playing a key role in the development of the illness phenotype, including psychotic symptomatology and cognitive impairments. There is emerging evidence for functional and structural hypoconnectivity between thalamus and prefrontal cortex in persons with schizophrenia spectrum disorders, as well as hyperconnectivity between thalamus and sensory and motor cortices. However, it is unclear whether thalamocortical dysconnectivity is a general marker of vulnerability to schizophrenia or a specific mechanism of schizophrenia pathophysiology. This study aimed to answer this question by using diffusion-weighted imaging to examine thalamocortical structural connectivity in 22 persons with schizophrenia or schizoaffective disorder (SZ), 20 siblings of individuals with a schizophrenia spectrum disorder (SIB), and 44 healthy controls (HC) of either sex. Probabilistic tractography was used to quantify structural connectivity between thalamus and six cortical regions of interest. Thalamocortical structural connectivity was compared among the three groups using cross-thalamic and voxel-wise approaches. Thalamo-prefrontal structural connectivity was reduced in both SZ and SIB relative to HC, while SZ and SIB did not differ from each other. Thalamo-motor structural connectivity was increased in SZ relative to SIB and HC, while SIB and HC did not differ from each other. Hemispheric differences also emerged in thalamic connectivity with motor, posterior parietal, and temporal cortices across all groups. The results support the hypothesis that altered thalamo-prefrontal structural connectivity is a general marker of vulnerability to schizophrenia, whereas altered connectivity between thalamus and motor cortex is related to illness expression or illness-related secondary factors.

## Introduction

Schizophrenia has been framed as a disorder of altered brain connectivity, with dysfunction in thalamocortical circuity argued to play a key role in the development of the illness phenotype (1). Various genetic and environmental risk factors could contribute to abnormal neurodevelopment, resulting in disruptions in neuronal connectivity and communication that ultimately manifest as impaired cognitive processes and clinical symptoms of schizophrenia (1). The thalamus occupies a significant position both anatomically and functionally that may mediate large-scale dysconnectivity: it relays sensory signals from subcortical regions to cortex and also actively participates in communication between different cortical areas, thus supporting basic sensorimotor coordination as well as higher-order cognitive functions like executive control and language (2, 3). Given its broad role in regulating cortical functioning, altered thalamocortical connectivity may thus provide a comprehensive neurological basis for the diverse clinical symptoms of schizophrenia, as well as the wide range of cognitive, social, and emotional impairments (4, 5) that are observed in the illness.

Increasing evidence from neuroimaging studies supports altered thalamocortical connectivity in persons with schizophrenia. The majority of this evidence comes from functional connectivity studies (see (6, 7) for review and (8) for meta-analysis), which report a consistent pattern of reduced coordination of resting endogenous activity between the thalamus and prefrontal cortex and increased coordination between thalamus and sensory and motor areas in individuals with schizophrenia, relative to healthy controls. Data from structural connectivity studies using diffusion-weighted imaging (DWI) and diffusion tensor imaging (DTI), though limited, revealed a similar pattern to functional connectivity studies: reduced connectivity between thalamus and prefrontal cortex (9–14) and, in a subset of the aforementioned studies, increased connectivity with sensory and motor areas (10–13). A similar thalamocortical connectivity pattern has also been found in individuals at clinical high risk for psychosis in both functional (15) and structural (12) imaging studies, which is furthermore predictive of conversion to a full-blown psychotic disorder (15) and associated with functional decline (12). In addition, this pattern of hypoconnectivity between thalamus and prefrontal cortex and hyperconnectivity between thalamus and sensorimotor regions has been observed in persons with psychotic bipolar disorder (13, 16–18), intimating the possibility that thalamocortical dysconnectivity may represent a transdiagnostic marker of psychosis. Taken together, these neuroimaging findings suggest that thalamocortical dysconnectivity may be a pathophysiological mechanism of schizophrenia, and possibly psychosis broadly.

It is still unclear, however, whether thalamocortical dysconnectivity in schizophrenia is a proximal illness mechanism or reflects (familial) vulnerability to develop the illness. Unaffected relatives of individuals with schizophrenia are at higher risk of the disorder due to their high proportion of shared genes (19) and common environment, and thus may provide insights into the significance of thalamocortical dysconnectivity in schizophrenia. Data from unaffected relatives are consistent with altered thalamic function, structure, and/or connectivity being a marker of vulnerability; however, interpretation of these findings is complicated somewhat by inclusion criteria. In some studies, relatives were screened only for history of psychotic disorder, whereas in others, relatives were screened for any psychiatric illness. Compared to healthy controls, previous studies have consistently found reduced working memory performance and altered fMRI activation in prefrontal cortex and bilateral thalamus during working memory tasks in healthy first-degree relatives with no psychiatric disorder (see (20) for a meta-analysis). The mediodorsal nucleus of thalamus is reciprocally connected to the prefrontal cortex, forming a key circuit supporting working memory maintenance (21). These results suggest that altered neural activation in thalamo-prefrontal circuits may indeed indicate a familial risk of schizophrenia. Other studies have found increased thalamic glutamate levels using magnetic resonance spectroscopy (22), reduced thalamic volume (see (23) for review), and reduced fractional anisotropy (the most widely used measure of microstructural alterations of white matter pathways with lower values indicating potential white matter neuropathology or normal aging) in the anterior limb of the internal capsule (24), superior longitudinal fasciculus (25), and the thalamus (26) in relatives screened for a history of psychotic disorder. Furthermore, several studies reported reduced thalamo-frontal functional connectivity in healthy siblings with no psychiatric illness (27, 28) and in relatives with no psychotic disorder (29, 30), but see (31). However, there were also important differences in the pattern of abnormal thalamocortical functional connectivity between unaffected relatives and participants with schizophrenia: in a recent study, increased sensorimotor-thalamic functional connectivity was observed in patients but not their unaffected siblings (28). To date, however, only one study has examined tractography-defined thalamocortical structural connectivity in first-degree relatives unaffected by psychosis and major mood disorders (32). Compared to healthy controls, relatives had reduced fractional anisotropy in the left thalamo-orbitofrontal tract. However, the interpretation of this finding is limited by the fact that the study did not examine the connectivity between thalamus and other cortical regions, nor were individuals with schizophrenia included as a comparison group.

In the current study, we sought to determine whether thalamocortical structural dysconnectivity, and thalamo-prefrontal hypoconnectivity more specifically, was a marker of familial risk towards schizophrenia or schizoaffective disorder. To this end, we investigated structural connectivity patterns between the thalamus and 6 cortical regions using probabilistic diffusion tensor tractography in individuals with schizophrenia or schizoaffective disorder, healthy siblings of individuals with a schizophrenia spectrum disorder, and healthy controls. We followed the analytic strategy of previous studies reporting altered structural thalamocortical connectivity in schizophrenia (11, 13), including the definition of specific cortical regions, in order to enable direct comparisons. We first expected to replicate previous findings of hypoconnectivity between thalamus and prefrontal cortex (9–14) and hyperconnectivity between thalamus and sensorimotor regions (10–13) in schizophrenia. Our primary research question was whether healthy siblings would show similar patterns of thalamocortical connectivity to individuals with schizophrenia, suggesting that these connectivity patterns are related to illness vulnerability, or whether they would look more similar to healthy controls, which would suggest that altered thalamocortical connectivity was related to illness expression or to other secondary factors related to schizophrenia (e.g. psychosocial consequences, antipsychotic medication use, etc.). Results of the current study will add to our understanding of the etiological significance of thalamocortical dysconnectivity in schizophrenia and in turn help the development of more targeted treatment and intervention strategies.

## Methods and Materials

### Participants

Eighty-seven participants between the ages of 18 and 55 completed this study. Twenty-two antipsychotic-medicated persons with schizophrenia or schizoaffective disorder (SZ) were recruited from both a longitudinal study (*n* = 19 participants who lived locally) (33) and an outpatient psychiatric facility (*n* = 3) in the Netherlands. Twenty healthy siblings of persons with a schizophrenia spectrum disorder (SIB) were recruited from the same longitudinal study.

Through community advertisements, 44 healthy individuals (HC) were recruited to serve as a control group. Diagnoses in SZ and SIB were established using Diagnostic and Statistical Manual of Mental Disorders, fourth edition (DSM-IV) criteria and verified with the Comprehensive Assessment of Symptoms and History interview (34) or Schedules for Clinical Assessment for Neuropsychiatry, version 2.1 (35). The explicit exclusion criteria for HC was a family history of psychotic disorders; however, none of the controls reported a first-degree relative with any psychiatric illnesses. Participants in the SIB and HC groups were excluded if they had any current Axis I disorder. Participants in all groups were excluded if they had a history of head trauma or neurological illness, or substance abuse or dependence within 6 months before the study. The participants in the SIB and SZ groups were not biologically related.

Demographic and clinical data are presented in **Table 1** and additional details on diagnoses are provided in Supplementary Methods. All SZ were taking antipsychotic medications, and chlorpromazine (CPZ) equivalent antipsychotic dosages were calculated (36). Clinical symptoms were assessed in SZ only with the Positive and Negative Syndrome Scale (PANSS) (37). Premorbid IQ was assessed with the Dutch version of the National Adult Reading Test (Nederlandse Leestest voor Volwassenen (38)). The Edinburgh Handedness Scale was used to measure handedness (39). The three groups did not differ significantly on sex, handedness, and IQ. SZ were significantly older than SIB and HC, so age was included as a covariate in all between-group analyses.

**Table 1.**
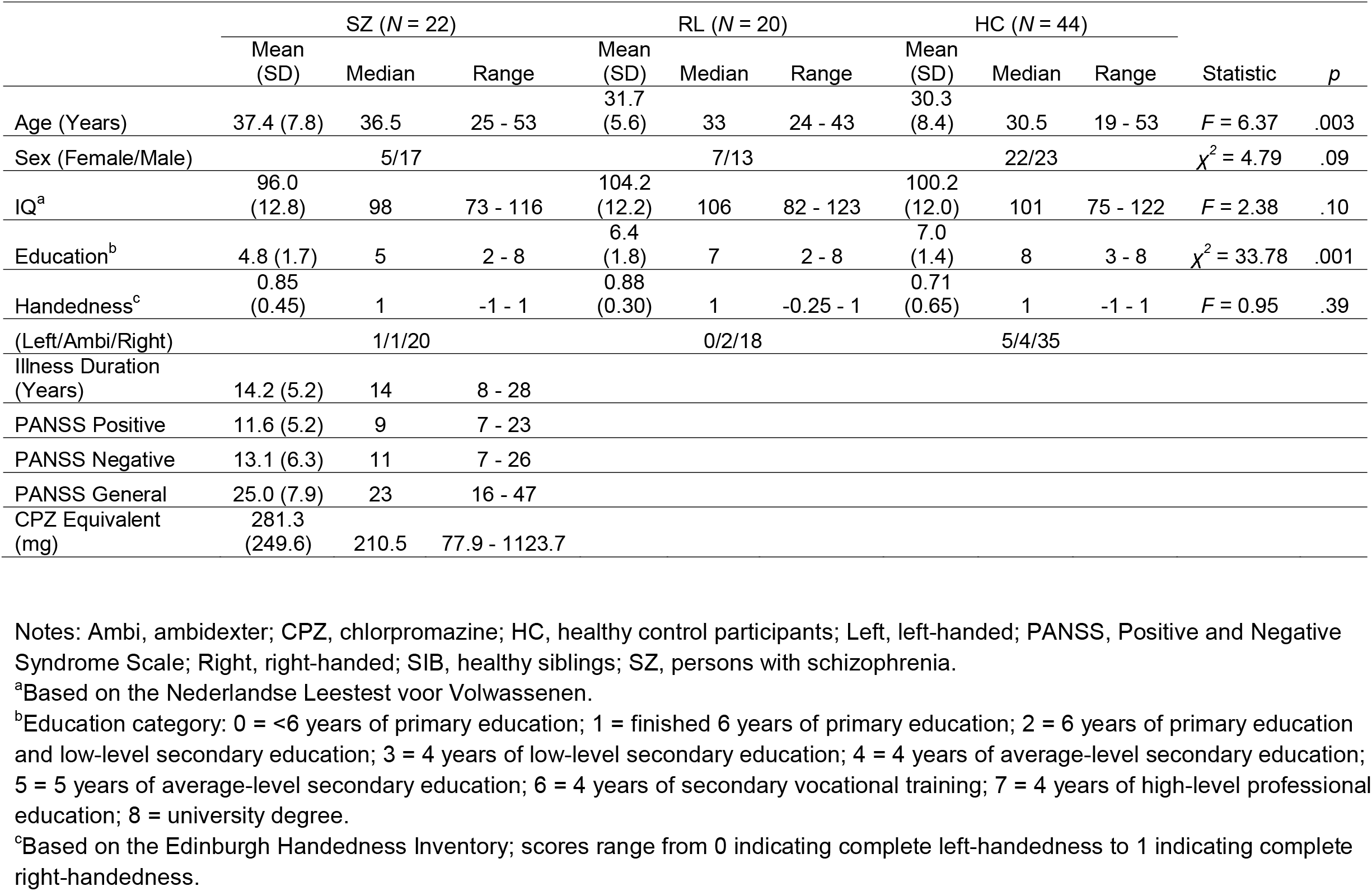
Demographic information.

|  | SZ (N = 22) |  |  | RL (N = 20) |  |  | HC (N = 44) |  |  | Statistic | p |
| --- | --- | --- | --- | --- | --- | --- | --- | --- | --- | --- | --- |
|  | Mean<br>(SD) | Median | Range | Mean<br>(SD) | Median | Range | Mean<br>(SD) | Median | Range |  |  |
| Age (Years) | 37.4 (7.8) | 36.5 | 25 - 53 | 31.7<br>(5.6) | 33 | 24 - 43 | 30.3<br>(8.4) | 30.5 | 19 - 53 | $F = 6.37$ | .003 |
| Sex (Female/Male) | | 5/17 | | | 7/13 | | | 22/23 | | $\chi^2 = 4.79$ | .09 |
| IQ<sup>a</sup> | 96.0<br>(12.8) | 98 | 73 - 116 | 104.2<br>(12.2) | 106 | 82 - 123 | 100.2<br>(12.0) | 101 | 75 - 122 | $F = 2.38$ | .10 |
| Education<sup>b</sup> | 4.8 (1.7) | 5 | 2 - 8 | 6.4<br>(1.8) | 7 | 2 - 8 | 7.0<br>(1.4) | 8 | 3 - 8 | $\chi^2 = 33.78$ | .001 |
| Handedness<sup>c</sup> | 0.85<br>(0.45) | 1 | -1 - 1 | 0.88<br>(0.30) | 1 | -0.25 - 1 | 0.71<br>(0.65) | 1 | -1 - 1 | $F = 0.95$ | .39 |
| (Left/Ambi/Right) |  | 1/1/20 |  |  | 0/2/18 |  |  | 5/4/35 |  |  |  |
| Illness Duration<br>(Years) | 14.2 (5.2) | 14 | 8 - 28 |  |  |  |  |  |  |  |  |
| PANSS Positive | 11.6 (5.2) | 9 | 7 - 23 |  |  |  |  |  |  |  |  |
| PANSS Negative | 13.1 (6.3) | 11 | 7 - 26 |  |  |  |  |  |  |  |  |
| PANSS General | 25.0 (7.9) | 23 | 16 - 47 |  |  |  |  |  |  |  |  |
| CPZ Equivalent<br>(mg) | 281.3<br>(249.6) | 210.5 | 77.9 - 1123.7 |  |  |  |  |  |  |  |  |

Notes: Ambi, ambidexter; CPZ, chlorpromazine; HC, healthy control participants; Left, left-handed; PANSS, Positive and Negative Syndrome Scale; Right, right-handed; SIB, healthy siblings; SZ, persons with schizophrenia.
<sup>a</sup>Based on the Nederlandse Leestest voor Volwassenen.
<sup>b</sup>Education category: 0 = <6 years of primary education; 1 = finished 6 years of primary education; 2 = 6 years of primary education and low-level secondary education; 3 = 4 years of low-level secondary education; 4 = 4 years of average-level secondary education; 5 = 5 years of average-level secondary education; 6 = 4 years of secondary vocational training; 7 = 4 years of high-level professional education; 8 = university degree.
<sup>c</sup>Based on the Edinburgh Handedness Inventory; scores range from 0 indicating complete left-handedness to 1 indicating complete right-handedness.

All participants gave written informed consent and were compensated for participation. The study was approved by the Medical Ethical Committee of the University Medical Center, Utrecht, The Netherlands. MRI data from a subset of current participants have been reported in previous studies (40–43).

### Diffusion-weighted imaging and probabilistic tractography

#### Image acquisition

All diffusion-weighted data were acquired at the University Medical Center Utrecht from 2012 – 2014 on an Achieva 3T scanner (Philips Medical Systems) equipped with an eight-channel head coil allowing parallel imaging. Two diffusion images were acquired using single-shot echoplanar imaging sequences, consisting of 30 diffusion-weighted scans (*b* = 1000 s/mm^2^) with noncollinear gradient directions and one image without diffusion weighting (*b* = 0 s/mm^2^), covering the entire brain [repetition time (TR) = 7057 ms; echo time (TE) = 68 ms; field of view = 240 × 240 × 150 mm; in-plane resolution = 1.875 × 1.875 mm; slice thickness = 2 mm; no slice gap; 75 axial slices; matrix size, 128 × 128 × 75]. The diffusion-weighted scans were measured twice, once with phase-encoding direction reversed (first scan, posterior-anterior; second scan, anterior-posterior), to correct for susceptibility-induced spatial distortions (44). For registration purposes, a whole-brain three-dimensional T1-weighted scan (200 slices; TR = 10 ms; TE = 4.6 ms; flip angle = 8°; field of view, 240 × 240 × 160 mm; voxel size: 0.75 × 0.8 × 0.75 mm) was acquired. Images were acquired roughly parallel to the anterior commissure – posterior commissure line (AC-PC line). The majority of the HC participants were scanned in 2013 while the SIB and SZ groups were scanned in 2014.

#### Preprocessing

Raw diffusion-weighted scans were first visually examined to ensure image quality and were subsequently preprocessed and analyzed using FDT within FSL 5.0 (FMRIB Software Library; www.fmrib.ox.ac.uk/fsl). As diffusion-weighted scans suffer from spatial distortions along the phase-encoding direction, two diffusion-weighted scans were acquired with reversed phase-encoding blips, resulting in pairs of images with distortions going in opposite directions. From these two images, the off-resonance field was estimated using a method similar to that described by (44) as implemented in FSL (45), with default cubic B-spline interpolation. Next, the 30 diffusion-weighted images from each phase-encoding direction were realigned to the *b*0 image using default affine registration procedures (12 parameters), and the *eddy_correct* command (with default settings such as trilinear interpolation) was used to correct for both eddy current induced distortions and subject movements simultaneously. The gradient directions were not rotated. No significant group differences in participant head movements were observed (see **Supplementary Methods**). The eddy-corrected scans with opposite phase-encoding blips were then combined into a single corrected image using the previously estimated off-resonance field with the *applytopup* command. The mean b0 image was extracted from the *topup* corrected volumes by using the *fslroi* command. A mask was then created from the mean b0 image and dilated twice using the *spm_dilate* function. The mask was then applied to all diffusion-weighted images.

#### Regions of interest

Each participant’s structural T1-weighted image was automatically segmented and labeled into known cortical and subcortical structures using Freesurfer 5.3 (http://surfer.nmr.mgh.harvard.edu). Segmentations were visually inspected, and no additional manual adjustments were needed. Selected cortical parcellations (**Table S1**) were combined to form six cortical regions of interest (ROIs) in each hemisphere: prefrontal cortex, motor cortex, somatosensory cortex, posterior parietal cortex, temporal cortex, and occipital cortex (**Figure 1**). To increase comparability, we followed previous studies as close as possible when choosing corresponding parcellations for specific ROIs (11, 13). The thalamus segmentation was then manually edited using MRIcron (https://www.nitrc.org/projects/mricron/) by the first author to include the lateral and medial geniculate nuclei and occasionally fill in caudal parts of the thalamus that were not captured by the Freesurfer segmentation, following an established protocol for manual segmentation of the human thalamus using bias-corrected T1-weighted images (46)(**Figure S1**). To test for any systematic group differences in the resulting thalamus masks, we conducted a supplementary analysis comparing the spatial location of voxels included in the thalamus masks among the groups (see **Supplementary Methods** and **Results**). Transformation matrices were derived for each participant by registering their mean *b*0 images to the Freesurfer conformed space using *bbregister* (47). The inverse of these registration matrices was then applied to transform the ROIs to diffusion space with trilinear interpolation. Each registration was visually inspected for quality control. When automated registration was not satisfactory, the transformation matrix was manually adjusted using the *tkregister2* tool and the adjusted registration matrix was applied to ROIs.

**Figure 1.**
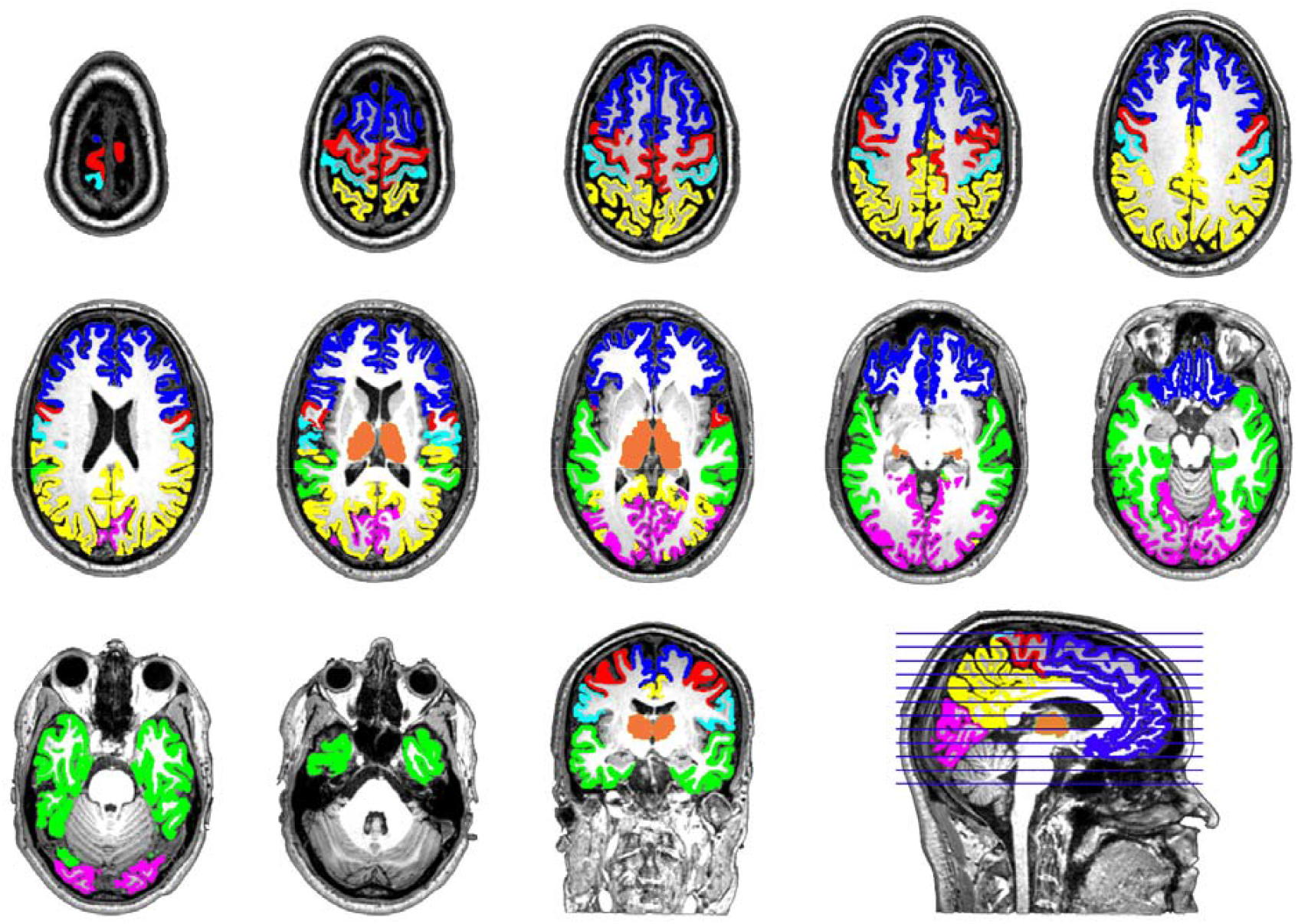
Cortical regions of interest (ROIs) and thalamus for a representative participant, displayed on the participant’s T1 image in horizontal view on multiple slices from superior to inferior. Masks based on Freesurfer’s automatic parcellation were combined to create an initial thalamus mask and six cortical ROI masks: prefrontal cortex (blue), motor cortex (red), somatosensory cortex (cyan), posterior parietal cortex (yellow), temporal cortex (green), and occipital cortex (magenta). The initial thalamus mask was then manually edited to include the lateral and medial geniculate nuclei to form the final thalamus mask (orange). These masks were then used in probabilistic tractography analyses to quantify thalamocortical connectivity.

#### Probabilistic tractography

Thalamocortical connectivity was assessed using probabilistic tractography, an analysis technique that reconstructs anatomical pathways between brain regions based on a distribution profile of probable fiber orientations in each voxel derived from diffusion-weighted images (48, 49). Within each hemisphere, probabilistic tractography was performed six times, using thalamus as the seed (start point) and one of the six cortical ROIs as a target (end point), using the *probtrackx2* command. The mid-sagittal plane was used as an exclusion mask (50–52), along with the other five cortical regions that were not the target at a given analysis, to avoid tracking continuing into the other hemisphere or passing through other cortical targets. The distribution profile of probabilistic connectivity was computed by sending out 5000 streamlines from each voxel within the thalamus, going in a direction drawn from a distribution around the principal diffusion direction until it was determined structurally impossible for a white matter tract to continue. Only streamlines that reached the target were preserved, and tracking stopped once a streamline reached the target. Pathways passing through the exclusion mask were rejected. Pathways that looped back on themselves were terminated, using the *loopcheck* option. Modified Euler integration as opposed to simple Euler for computing probabilistic streamlines was used to increase accuracy. Distance correction was used so that the connectivity distribution is the expected length of the pathways that cross each voxel times the number of streamlines that cross it, in order to correct for the fact that the number of streamlines drops with distance from the seed mask. All other settings were kept at default (e.g., curvature threshold: 0.2, number of steps: 2000, step length: 0.5, and fiber volume threshold: 0.01). Final tractography results for each individual were visually inspected for quality assurance. Two crossing fibers per voxel were modeled. Previous research shows that with 30 diffusion directions and diffusion-weighting *b* = 1000 s/mm^2^, two crossing fibers with a minimum angle of 50 degree can be reliably identified with a smaller than 20% false negative rate (53). From this analysis, six seed-to-target images were generated within both hemispheres where each voxel in the thalamus contains a value representing the number of streamlines originating from that voxel that reached the corresponding cortical ROI target.

#### Cross-thalamic cortical connectivity analysis

To increase comparability, we calculated our main measures of interest the same way as previous studies did (10–12). To examine group differences in the connectivity pattern between thalamus and each cortical ROI within each hemisphere, we computed percent connectivity for each thalamus-ROI pair as follows:

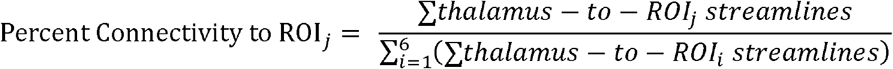

The absolute connectivity between thalamus and each cortical ROI was quantified as the sum of streamlines originating across all voxels in the thalamus that reached the corresponding ROI (i.e. the sum of values for each seed-to-target image). Total thalamocortical connectivity for a particular hemisphere was calculated as the sum of all streamlines from the thalamus that reached any of the six cortical ROIs (i.e. the sum of values for each seed-to-target image summed across all six seed-to-target images). To control for individual differences in total thalamocortical connectivity, percent connectivity was calculated for each cortical ROI by dividing its absolute connectivity with thalamus by total thalamocortical connectivity for that hemisphere (10). These tractography-defined connectivity values were measures of relative connectivity that were minimally biased by individual differences in brain size and global white matter fractional anisotropy. In other words, these relative connectivity values are protected against factors that could systematically affect the *absolute* number of streamlines reaching cortical targets. They were also independent of where the tract originated from inside the thalamus and are closer to a summary measure of structural connectivity patterns rather than a diffusion measure (e.g., fractional anisotropy) of any specific white matter tract. These percent connectivity measures were used as the dependent variables in main group comparisons.

#### Voxel-wise thalamocortical connectivity analysis

To localize putative group differences in thalamocortical connectivity patterns, we computed a probability value at each thalamic voxel for each cortical ROI as follows:

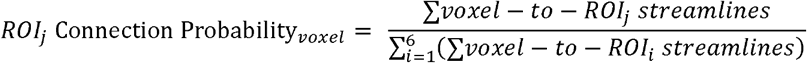

From each seed-to-target image, probability maps were calculated by dividing the value at each voxel (representing the number of streamlines that arrived at a particular cortical ROI) by the sum of all six images at each voxel. These probability maps thus represent the probability of a given voxel in the thalamus connecting with a particular cortical ROI, relative to general thalamocortical connectivity.

In order to examine voxel-wise thalamocortical connectivity differences across groups, we normalized individual probability maps. Specifically, each participant’s anatomical T1-weighted volume was realigned to their mean *b*0-weighted image and subsequently segmented into gray matter, white matter, and CSF, and normalized to MNI space using the unified segmentation algorithm as implemented in SPM8 (54). Then all probability maps were transformed into MNI space using each individual’s normalization matrix, and then averaged within each group. To exclude voxels that don’t contain reliable projections to cortex, we adopted a procedure used in previous papers (11, 13). First, we created a within-group mask for each group to include all voxels in the thalamus that have a value >10% for any of the cortical ROI targets. The union of these three group masks were then binarized to form the final inclusion mask that contained all voxels with a value >10% for any group. Normalized individual probability maps were smoothed using a 4mm kernel before the inclusion mask was applied. These normalized, smoothed, and masked probability maps were then analyzed in voxel-wise group comparisons using SPM12 (http://www.fil.ion.ucl.ac.uk/spm/software/spm12).

### Statistical analysis

#### Demographic comparisons

Groups were compared using one-way ANOVAs on age, IQ, and handedness. Chi-squared tests were employed to compare groups on sex and education.

#### Cross-thalamic cortical connectivity analysis

A careful examination of the Q-Q plots revealed that many of the connectivity values were not normally distributed. Therefore, a logit transformation was applied to these connectivity values in order to transform the distribution to normal and stabilize the variance (55). After transformation, Q-Q plots were visually examined again to ensure normality was achieved. Box’s M test revealed that the assumption of homoscedasticity was met after transformation (*M* = 211.68, *p* = .41).

To protect against inflated Type I error due to multiple comparisons (56), a MANCOVA was conducted in SPSS Statistics version 25.0 (IBM) to examine if there were overall group differences across the six measures of percent connectivity between each cortical ROI and the thalamus. Diagnostic group was included as a between-subject variable and age as a covariate. Hemisphere was included as a within-subject variable due to its understudied potential significance in thalamocortical connectivity (57). Significant main effects were followed up by univariate tests: individual group comparisons of percent connectivity between each cortical ROI and the thalamus were evaluated with a repeated-measures ANCOVA, with diagnostic group as a between-subject variable, hemisphere as a within-subject variable, and age as a covariate. Significant main effects of group were then followed up with pairwise comparisons, using Sidak adjustment (very similar to Bonferroni correction) for multiple comparisons (58), and accompanied by Cohen’s *d* to quantify effect sizes. Though sex was not significantly different among groups, we conducted a supplementary analysis including sex as a covariate in the analysis detailed above. To investigate the possibility of any potential connectivity differences due to differences in ROI size, we conducted a supplementary analysis comparing total volumes of thalamus and cortical ROI among the groups (see **Supplementary Methods** and **Results**). To examine the potential confounding effect of antipsychotic use, CPZ equivalent dose (36) was correlated with total percent connectivity using Spearman’s rank correlation (*r*_s_).

#### Voxel-wise thalamocortical connectivity analysis

For each cortical ROI within each hemisphere, ANCOVAs were conducted at each voxel in the seed-to-target probability maps, with diagnostic group as a between-subject variable and age as a covariate. Statistical maps were tested for a significant effect of group using cluster-level inference (i.e., the inference that the cluster of voxels driving the effect is highly unlikely to have occurred by chance; cluster-defining threshold at voxel level, *p* < .001; cluster probability of *p* < .05, family wise error-corrected for multiple comparisons).

## Results

### Cross-thalamic cortical connectivity

Percent connectivity between each cortical ROI and the thalamus in all three groups are presented in **Figure 2**. Across all six cortical ROIs, the MANCOVA test revealed significant main effects of group (*F*(12,154) = 1.90, *p* = .04; Wilk’s Λ = 0.76, partial *η*^*2*^ = 0.13) and hemisphere (*F*(6,78) = 8.91, *p* < .001; Wilk’s Λ = 0.59, partial *η*^*2*^ = 0.41). There was no significant group × hemisphere interaction effect (*F*(12,156) = 1.12, *p* = .35; Wilk’s Λ = 0.85, partial *η*^*2*^ = 0.08), nor any effect of age (*F*(6,77) = 0.89, *p* = .51; Wilk’s Λ = 0.94, partial *η*^*2*^ = 0.07).

**Figure 2.**
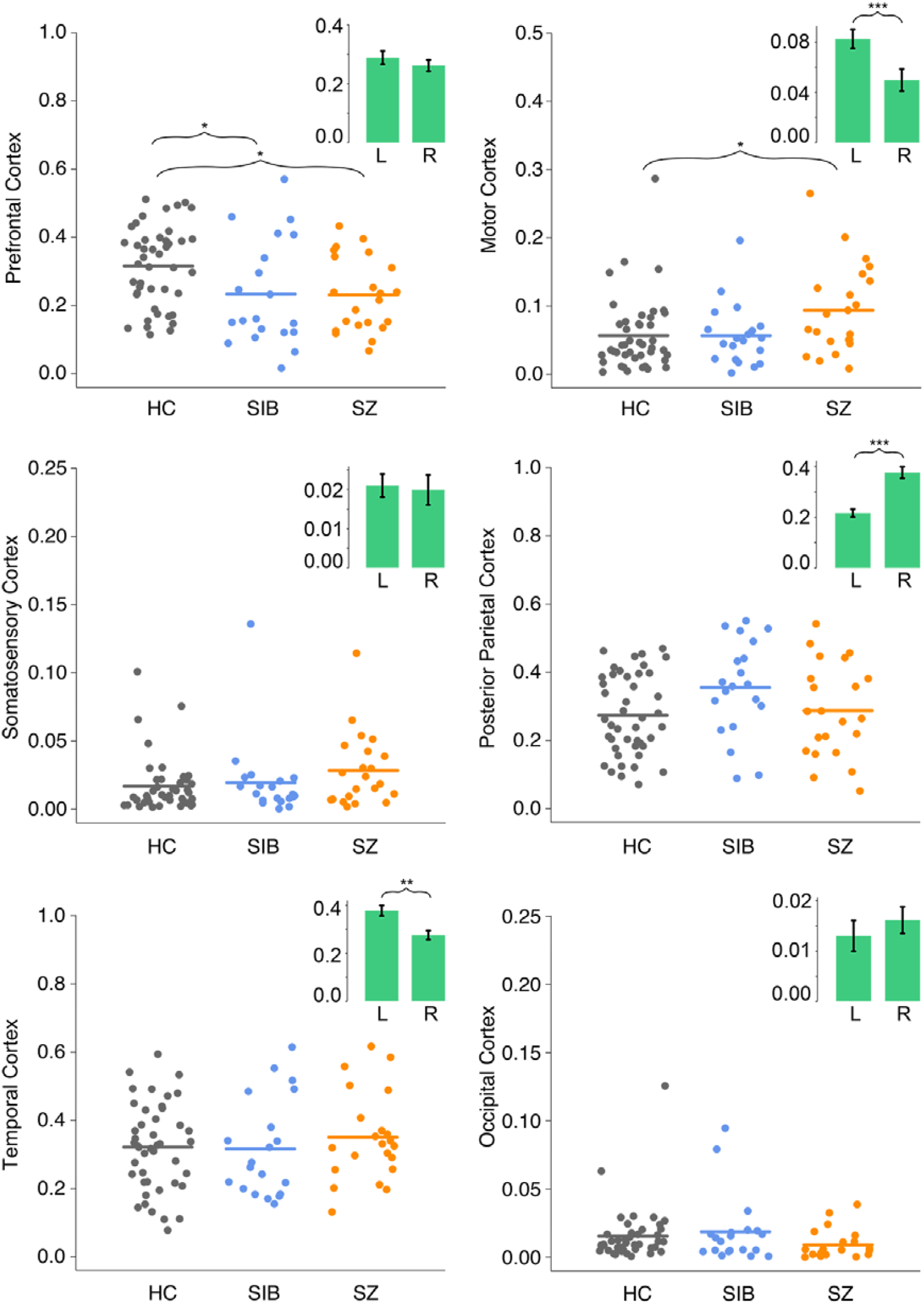
Structural connectivity between cortical regions and thalamus for each group collapsed across hemispheres. Points represent percent connectivity values before logit transformation. Insets show hemisphere effects collapsed across groups. HC, healthy control participants; L, left hemisphere; SIB, healthy siblings; SZ, persons with schizophrenia; R, right hemisphere. * *p* < .05, ** *p* < .01, *** *p* < .001.

For connections between thalamus and prefrontal cortex, there was a significant main effect of group (*F*(2,82) = 6.17, *p* = .003, partial *η*^*2*^ = 0.13). Post-hoc pairwise comparisons after Sidak adjustment revealed that, compared to HC, both SIB (*p* = .02, Cohen’s *d* = 0.8) and SZ (*p* = .02, Cohen’s *d* = 0.8) had significantly lower thalamo-prefrontal connectivity. SIB and SZ did not differ from each other (*p* = .99, Cohen’s *d* = 0.04). There was no significant effect of hemisphere (*F*(1,83) = 0.78, *p* = .38, partial *η*^*2*^ = 0.01), nor any group by hemisphere interaction (*F*(2,83) = 1.29, *p* = .28, partial *η*^*2*^ = 0.03). For connections between thalamus and motor cortex, there was also a significant effect of group (*F*(2,82) = 4.47, *p* = .01, partial *η*^*2*^ = 0.10). Post-hoc tests with Sidak adjustment revealed significantly higher connectivity in the SZ than HC group (*p* = .01, Cohen’s *d* = 0.8), while SIB did not differ significantly from either HC (*p* = .95, Cohen’s *d* = 0.1) or SZ (*p* = .10, Cohen’s *d* = 0.7). There was also a significant main effect of hemisphere (*F*(1,83) = 22.90, *p* < .001, partial *η*^*2*^ = 0.22), with thalamo-motor connectivity being higher in the left hemisphere, but no significant group-by-hemisphere interaction effect (*F*(2,83) = 0.31, *p* = .73, partial *η*^*2*^ = 0.01).

There was no group difference in percent connectivity between thalamus and any other cortical ROI (Somatosensory: *F*(2,82) = 2.71, *p* = .07, partial *η*^*2*^ = 0.06; Posterior Parietal: *F*(2,82) = 2.23, *p* = .11, partial *η*^*2*^ = 0.05; Temporal: *F*(2,82) = 0.59, *p* = .56, partial *η*^*2*^ = 0.01; Occipital: *F*(2,82) = 1.96, *p* = .15, partial *η*^*2*^ = 0.05). There was, however, a significant effect of hemisphere on thalamo-posterior parietal (*F*(1,83) = 27.73, *p* < .001, partial *η*^*2*^ = 0.25) and thalamo-temporal (*F*(1,83) = 11.07, *p* = .001, partial *η*^*2*^ = 0.12) connectivity. Thalamo-posterior parietal connectivity was higher in the right hemisphere, while thalamo-temporal connectivity was higher in the left hemisphere. There was no hemisphere difference in percent connectivity between thalamus and somatosensory cortex (*F*(1,83) = 2.60, *p* = .11, partial *η*^*2*^ = 0.03) and between thalamus and occipital cortex (*F*(1,83) = 1.99, *p* = .16, partial *η*^*2*^ = 0.02). Lastly, there was no significant group × hemisphere interaction effect on connectivity between thalamus and either somatosensory (*F*(2,83) = 2.88, *p* = .06, partial *η*^*2*^ = 0.06), posterior parietal (*F*(2,83) = 0.29, *p* = .75, partial *η*^*2*^ = 0.01), temporal (*F*(2,83) = 2.37, *p* = .10, partial *η*^*2*^ = 0.05), or occipital (*F*(2,83) = 1.07, *p* = .35, partial *η*^*2*^ = 0.03) cortex.

There was also no effect of age on connectivity between thalamus and any of the cortical ROIs (.07 ≤ *p* ≤ .91). Results were similar when including sex as a covariate (see **Supplementary Results**). Additionally, there were no correlations between antipsychotic dosage and thalamocortical connectivity for any of the cortical ROIs (-.27 ≤ *r*_*s*_ ≤ .32, .20 ≤ *p* ≤ .99). Finally, there were no group differences in total volumes of thalamus and cortical ROI or in the voxel composition of thalamus masks used in the probabilistic tractography analysis (**Table S2**).

### Voxel-wise thalamocortical connectivity

Voxel-wise connectivity patterns were qualitatively similar across all groups (**Figure S2**). However, there were no clusters in which thalamocortical connectivity differed significantly between groups after correcting for multiple comparisons. For the purpose of aiding future meta-analyses and comparison with other studies, we reported clusters significant at *p* < .005 after applying an extent threshold of ≥ 10 voxels (see **Table S3** & **Table S4**), following published guidelines (59, 60).

## Discussion

In this study, we used probabilistic tractography analyses of DWI data to examine thalamocortical structural connectivity patterns in persons with schizophrenia and schizoaffective disorder (henceforth referred to as persons with schizophrenia), healthy siblings of persons with a schizophrenia spectrum disorder, and healthy controls. We found reduced connectivity between thalamus and prefrontal cortex in both persons with schizophrenia and healthy siblings, compared to healthy controls. In addition, compared to healthy controls, persons with schizophrenia had increased structural connectivity between thalamus and motor cortex; healthy siblings did not. Moreover, we observed hemispheric differences in several thalamocortical connectivity patterns across all groups. Taken together, these findings are consistent with prior reports in schizophrenia, and results from healthy siblings suggest that hypo-connectivity between thalamus and prefrontal cortex may represent a marker of vulnerability related to familial risk. In this way, these data thus provide novel insights into the etiology and functional significance of thalamocortical dysconnectivity in the illness.

Using largely identical analytic methods to previous studies, we replicated findings of hypoconnectivity between thalamus and prefrontal cortex in persons with schizophrenia. These findings converge with other reports of reduced structural (9–14) and functional (see (6) for review) thalamo-frontal connectivity. We also identified a hyperconnectivity between thalamus and motor cortex in persons with schizophrenia. Findings of structural and functional connectivity between thalamus and cortical regions other than prefrontal cortex are less consistent. While Marenco and colleagues (10) found elevated structural connectivity between thalamus and somato-motor cortex - consistent with our own findings - other studies did not, instead reporting increased thalamic connectivity with somatosensory (11, 13), occipital (11), and parietal (12) cortices. The hyperconnectivty between thalamus and motor cortex is, however, in line with functional connectivity findings (see (6) for review). The inconsistent findings regarding hyper-thalamocortical connectivity may be due to differences in imaging parameters, methods of defining cortical ROIs, clinical heterogeneity, and statistical power across studies.

More importantly, our study showed reduced thalamo-prefrontal connectivity in healthy siblings of persons with schizophrenia, compared to healthy controls, corroborating recent findings of reduced fractional anisotropy between thalamus and orbitofrontal cortex in first-degree relatives unaffected by psychosis and major mood disorders (32). The degree of thalamo-prefrontal hypoconnectivity did not differ between healthy siblings and persons with schizophrenia. The combined pattern of reduced thalamo-prefrontal connectivity and non-elevated thalamo-motor connectivity in healthy siblings mirrors recent functional connectivity findings in a large sample of individuals with schizophrenia, unaffected siblings, and healthy controls (28). Given that siblings were screened for any current mental illness and had, by and large, passed the age by which a psychotic disorder typically emerges, these results suggest that thalamo-prefrontal hypoconnectivity is neither a proximal mechanism of schizophrenia symptomology nor secondary to chronic antipsychotic use or the psychosocial consequences of having a severe mental illness. Instead, our results suggest that it is a correlate of familial risk towards schizophrenia.

Naturally, this finding in healthy siblings draws into question the functional significance of altered thalamo-prefrontal connectivity. Siblings had equivalent reductions in connectivity between the thalamus and prefrontal cortex, but did not exhibit clinically significant symptoms. One simple explanation is that thalamo-prefrontal hypoconnectivity does not contribute to the emergence of schizophrenia symptoms. However, this seems unlikely given previous findings in persons at clinical high risk of developing a psychotic disorder: thalamocortical functional dysconnectivity is predictive of conversion to a full-blown psychotic disorder (15) and reduced thalamo-orbitofrontal structural connectivity is associated with functional decline (12). These data would suggest that thalamo-prefrontal hypoconnectivity may indeed contribute to the onset of formal psychosis.

Another explanation is the presence of protective factors in healthy siblings. In considering this question, we revisit the function of thalamocortical connections and the implications of dysfunction in these circuits. The thalamus is a central hub in the brain that not only relays basic sensory information from subcortical structures to the cortex, but also participates in almost all intracortical communications (2). The functional specialization and organization of thalamic nuclei and the formation of thalamocortical circuits involve complicated interactions between the thalamus and the cortex through gene expressions and neuronal signaling (61, 62). Misexpression of certain genes can lead to thalamic rearrangements through rewiring of thalamocortical axons or changes in thalamic structures (61). Consequently, alterations in this neurodevelopmental process could lead to atypical connection patterns between thalamus and different cortical regions. Indeed, in functional connectivity studies, thalamo-prefrontal hypoconnectivity and thalamo-sensory/motor hyperconnectivity in persons with schizophrenia are inversely correlated (see (8) for a meta-analysis), suggesting a potential common underlying mechanism. It is possible that the formation of thalamocortical circuits during development is altered in individuals with a familial liability towards schizophrenia, but to a lesser degree in healthy siblings. That is, the wiring of thalamocortical circuits may be more imbalanced in persons with schizophrenia and give rise to both hypoconnectivity and hyperconnectivity, but only hypoconnectivity in healthy siblings. The lack of thalamo-motor hyperconnectivity may in turn serve as a protective factor for healthy siblings such that the weak top-down control (as evidenced by thalamo-prefrontal hypoconnectivity) was not further exacerbated by aberrant sensorimotor control. This possibility fits neatly with recent computational accounts of psychosis that posit abnormal weighting of top-down expectations and incoming sensory information as a potential disease mechanism (63, 64), where thalamus potentially plays a crucial role in relaying expectations and sensory afferents (2, 65). However, exactly what genetic and environmental input could serve as protective factors during key stages of thalamocortical circuit development is still a question for future research (61).

Our study also identified hemispheric differences in thalamocortical structural connectivity. Across all groups, connectivity between thalamus and both motor and temporal cortex was higher in the left hemisphere and connectivity between the thalamus and posterior parietal cortex was higher in the right hemisphere. The higher left thalamo-motor connectivity may reflect the fact that a large proportion of participants across all groups are right-handed, as right-handedness is associated with larger volume and higher fractional anisotropy in the left relative to the right motor cortex (66, 67). Higher left thalamo-temporal connectivity is consistent with findings of larger white matter volumes in the left relative to the right primary auditory cortex (68), and mirrors findings in cortico-cortical white matter pathways: arcuate fasciculus (a major frontal-temporal association fiber tract) has larger tract volume, higher relative fiber density, and higher fractional anisotropy in the left relative to the right hemisphere (69–72). Functional imaging data also consistently shows larger activation during language tasks in the left relative to the right thalamus (73), which may be supported by a higher thalamo-temporal structural connectivity in the left hemisphere. Similarly, higher right thalamo-posterior parietal connectivity mirrors findings of larger tract volume and higher fractional anisotropy in the right relative to the left superior longitudinal fasciculus (a major association fiber bundle connecting the frontal and posterior parietal cortices) (74, 75).

Interpretation of the current findings is limited by several factors. First, the results from our cross-thalamic analysis did not align with the results from the voxel-wise analysis, which did not reveal any regions where thalamo-cortical connectivity differed between groups after correction for multiple comparisons. One possible explanation is large individual differences in the topographic organization of thalamic nuclei (76), which would reduce overlap in peak connectivity values across participants and lead to reduced signal after group averaging. The cross-thalamic approach circumvents this problem, thereby increasing statistical power, by averaging within participants. Precise spatial localization of thalamo-cortical connections may have also been limited by our number of diffusion gradient directions. Using probabilistic tractography, previous studies with more gradient directions were able to render sub-thalamic divisions close to histological segmentation results (77, 78). Though 30 gradient directions has been found to yield reliable tractography results for robust anatomical pathways (79–81) that do not differ substantially from 120 gradient directions (53), with fewer gradient directions, there is a greater chance of false negatives for fiber identification if the angle at which fibers cross is small (53). Thus, the voxel-wise analysis results may not reflect the *precise* functional subdivisions of the thalamus, leading to a more spatially diffuse pattern of group differences. Providing credence to these findings despite aforementioned methodological limitations is our replication of previous results in individuals with schizophrenia using identical cross-thalamic cortical connectivity analyses (10–13) and the striking overlap with findings of a recent functional connectivity study reporting shared reductions in thalamo-frontal connectivity in individuals with schizophrenia and unaffected siblings and increases in thalamo-motor connectivity in patients only (28). A second limitation is that tractography can only infer the direction of white matter tracts and cannot compute results at the level of individual axons.

Consequently, the technique may produce erroneous results when fibers cross or branch within a single voxel or when there is no dominant direction of water diffusion (50). Even though DWI-based tractography results of major white matter tracts have been repeatedly validated by postmortem dissections, the results of this study should not be interpreted as direct visualization of anatomical pathways. Similarly, the percent connectivity values likely describe an average over multiple white matter routes between the thalamus and cortical ROIs, particularly for the larger cortical regions. Accordingly, group differences in more localized tracts may be washed out in the current analyses. Future studies with larger samples and more diffusion directions may reveal potential group differences in structural connectivity between thalamus and smaller cortical subdivisions. Third, we were not able to separate contributions of familial environment versus genes in conferring risk towards schizophrenia. Future studies on healthy twins of persons with schizophrenia are needed to tease out the unique contribution of genes to thalamocortical structural connectivity abnormalities. Fourth, the thalamus mask generated by Freesurfer segmentation may have included non-thalamic tissue. This could have contributed to small inaccuracies in the derived percent connectivity measures. However, given that there were no systematic differences in voxel composition of the thalamus masks, we do not expect these small inaccuracies to have biased the groups effects. Lastly, healthy controls were, for the most part, scanned prior to individuals with schizophrenia and unaffected siblings, which raises the possibility of group differences being confounded with changes in scanner performance over time.

In conclusion, we identified significant decreases in thalamo-prefrontal structural connectivity in healthy siblings of persons with schizophrenia relative to healthy controls, which was not distinguishable from that in persons with schizophrenia. This finding suggests that thalamo-prefrontal hypoconnectivity may be a marker of familial vulnerability to schizophrenia and has important implications for understanding disease mechanisms and, thus, treatment development.

## Data Availability

Data is available upon request.

## Acknowledgements

This work was supported by the National Institutes of Health Grant R01-MH-112644 (K.N.T.), National Institute of Mental Health Grant R21-MH-115297-01 (K.N.T.), a NARSAD Young Investigator Award from the Brain and Behavior Foundation (K.N.T.), a Netherlands Organization for Scientific Research Rubicon grant (K.N.T.), a short-stay fellowship from Utrecht University (K.N.T.), and a University of Utrecht Neuroscience and Cognition grant (S.F.W.N.). The results from a preliminary analysis of this study was previously presented as a poster at the 32^nd^ Annual Meeting of the Society for Research in Psychopathology (SRP), Indianapolis, IN. The authors would like to thank Dr. Neil Woodward for his guidance on manual thalamus segmentation.

